# The effect of heat events on prehospital and retrieval service utilisation in rural and remote areas: a scoping review

**DOI:** 10.1101/2021.04.12.21255254

**Authors:** Elen O’Donnell, Bridget Honan, Simon Quilty, Rebecca Schultz

## Abstract

**Introduction:** It is well established that heatwaves increase demand for emergency transport in metropolitan areas, however little is known about the impact of heat events on demand for prehospital retrieval services in rural and remote areas, or how heatwaves are defined in this context.

**Inclusion criteria:** Papers were eligible for inclusion if they reported on the impact of a heat event on the activity of a prehospital and retrieval service in a rural or remote area.

**Methods:** A search of PubMed, Cochrane, Science Direct, CINAHL and Google Scholar databases was undertaken on 18 August 2020, using search terms related to emergency medical transport, extreme heat and rural or remote. Data relevant to the impact of heat on retrieval service activity was extracted, as well as definitions of extreme heat.

**Results:** Two papers were identified, both from Australia. Both found that heat events increased the number of road ambulance call-outs. Both studies used the Excess Heat Factor to define heatwave periods of interest.

**Conclusions:** This review found almost no primary literature on demand for prehospital retrieval services in rural and remote areas, and no data specifically related to aeromedical transport. The research did recognise the disproportionate impact of heat-related increase in service demand on rural and regional health services. With the effects of climate change already being felt, there is an urgent need for more research and action in this area.

## Introduction

### Background

The impact of extreme heat events on health is well documented across a range of settings. In Australia, heatwaves are responsible for more deaths than all other climate related disasters combined(1). Extreme heat events are associated with increased health service utilisation, including emergency department visits, hospital inpatient admissions, and urban ambulance call-outs (2-4).

There is no consensus definition of an extreme heat event. The Australian Bureau of Meteorology (BoM) defines a heatwave as a three-day period where both maximum and minimum temperatures are unusually hot compared to the normal local climatic conditions(5). However, in many parts of Australia, particularly northern Australia, heat is seasonal and extreme temperatures can last months.

Severe heat exposure can cause morbidity and mortality directly via heat-related illness, or more commonly indirectly, through exacerbation of underlying chronic disease. Those with medical comorbidities, particularly cardiorespiratory(6), renal(7) and mental illness(8) are at increased risk of morbidity and mortality during extreme heat events.

Natural tolerance for environmental heat falls within a narrow temperature range. Adaptive responses to heat include physiological, behavioural and sociocultural interventions including infrastructure such as the provision of environmentally cool spaces. Due to poor quality housing and energy insecurity, environmental cooling is not an option for everyone, contributing to the disproportionate effect of extreme heat events on people suffering socioeconomic disadvantage(9).

Most literature addressing the links between heat and health is focused on urban environments. Rural and remote environments present different challenges in a number of ways. Firstly, rural and remote communities frequently have increased rates of chronic disease and socioeconomic disadvantage compared to their urban counterparts, making these populations potentially more vulnerable to extreme heat events. Secondly, rural and remote health systems often have limited resources, and thus may be overwhelmed by even a small increase in demand during heat events. Lastly, the impact of extreme heat events shows geographical and climatic variation(10) with higher health burdens experienced in hotter climates where there is greater extent of hot weather (11). While there is a recognised “urban heat island effect” which tends to make densely populated urban areas warmer than the surrounding countryside, many major urban centres in Australia are located in temperate, coastal regions. Rural and remote regions are spread across the full range of climatic zones in Australia, including hot arid and tropical areas where baseline temperatures already approach the limits of human physiological adaptation, meaning that small increases may be more keenly felt.

### Importance

Extreme heat events are already increasing in severity, duration and frequency due to anthropogenic climate change (12). This trend is expected to continue even in a best-case scenario of emissions reductions, but if emissions continue to rise in line with current trends, unusually extreme heat events will become a yearly phenomenon. Therefore, there is an imperative for health services to urgently undertake further research on how heatwaves will impact on service delivery.

### Objective

Little is known about the impact of heatwaves on demand for prehospital and retrieval services in rural and remote areas. Furthermore, there is no consensus on definitions of rurality or heatwaves in the literature. The purpose of this study was to identify gaps in current knowledge and understanding of the impact of extreme heat events on demand for prehospital and retrieval services in rural and remote areas and clarify how rurality and heatwaves are defined in the literature. The objective of this literature review is to identify deficit areas for future research, and to facilitate policy-making for prehospital and retrieval services in rural and remote areas.

## Methods

A scoping review methodology was chosen in order to define the scope of published literature on the topic and identify gaps in current knowledge to inform future research, as well as explore concepts related to definitions of rurality and heatwaves as they relate to prehospital and retrieval services (13).

### Eligibility criteria

The research question was developed using the Population, Exposure, Comparator, Outcome framework to develop the search strategy. The population was patients of any age who required prehospital and retrieval services in a rural or remote region. Limiting the study to rural regions was important to differentiate from the previously well-established effects on urban regions. There is no consensus definition of rurality, therefore the investigators accepted any population described within the paper as rural or remote, taking into consideration population density and size, geographical distance from major centres, access to health care, and cited definitions of rurality (14, 15). The exposure was an extreme heat weather event, such as a heatwave. There is no consensus in the literature on the definition, therefore the investigators accepted any definition of extreme heat or heatwave event that was described within the paper. The comparator was non-heatwave periods. The outcome of interest was activity of a prehospital and retrieval service. This could relate to the frequency, acuity, complexity or urgency of the tasks.

### Search strategy

An initial limited search of PubMed was undertaken to identify articles on the topic. The text words contained in the titles and abstracts of relevant articles, and the index terms used to describe the articles were used to develop a full search strategy for PubMed, Cochrane, Science Direct, CINAHL and Google Scholar (see Table 1). The search included English-language papers published from January 2000 through October 2020. A secondary hand search of bibliographies was conducted to identify further outputs.

**Table 1.** Search Strategy

| Database | Search terms |
| --- | --- |
| PubMed | ((emergency medical services[MeSH Major Topic]) OR ("air medical"[Title/Abstract] OR aeromedical[Title/Abstract])) AND ((extreme heat[MeSH Major Topic]) OR (heat*[Title/Abstract] OR weather[Title/Abstract] OR climate[Title/Abstract]))<br>*limited to 10 years and English language |
| Cochrane | ((emergency medical services[MeSH Major Topic]) OR ("air medical" OR aeromedical)) AND ((extreme heat[MeSH Major Topic]) OR (heat* OR weather OR climate))<br>*limited to 10 years and English language |
| Science direct | (heat OR weather OR climate) AND ("medical retrieval" OR "air medical" OR aeromedical OR "emergency transport" OR ambulance)<br>*limited to Review articles and Research articles |
| CINAHL | (MH extreme heat OR AB ( heat* OR weather OR climate )) AND (MH emergency medical services OR AB ( "air medical" OR aeromedical )) |
| Google scholar | (heat OR weather OR climate) AND ("medical retrieval" OR "air medical" OR aeromedical OR "emergency transport" OR ambulance)<br>*limited to 100 most relevant |

### Inclusion Criteria

All peer-reviewed studies of the impact of heat wave events on prehospital and retrieval service activity in a rural or remote setting, in a high-income country, were included.

### Exclusion Criteria

Literature published in a language other than English, abstracts, citations, thesis, news media reports, case reports and reports unrelated to heatwaves or prehospital and retrieval service activity were excluded. Studies from low or middle-income countries, or based in metropolitan or urban settings, were also excluded.

### Data collection

Following the search, all identified citations were collated and uploaded into EndNote(16) and duplicates removed. Following a pilot test, titles and abstracts were screened by two independent reviewers for assessment against the inclusion criteria for the review. Potentially relevant sources were retrieved in full and their citation details imported into a spreadsheet. The full text of selected citations were assessed in detail against the inclusion criteria by two independent reviewers. Reasons for exclusion of sources of evidence at full text that did not meet the inclusion criteria were recorded and reported. Any disagreements that arose between the reviewers at each stage of the selection process were resolved through discussion. The results of the search and the study inclusion process are presented in a Preferred Reporting Items for Systematic Reviews and Meta-analyses extension for scoping review (PRISMA-ScR) flow diagram(17).

### Data Extraction

Data was extracted from full texts by two reviewers independently using a data extraction tool developed by the research team. The data extracted included specific details about the setting, service (description of the prehospital and retrieval service), population, exposure (how heatwave was defined), the outcome measure (how the utilisation of the prehospital retrieval service was measured) and the key results from the study.

### Synthesis of results

No assessment of bias was undertaken due to being a scoping review. Results were summarised in a table and narrative form.

## Results

The search strategy yielded a total of 793 references. After exclusion of duplicates, 739 titles and abstracts were identified for further screening. After applying exclusion criteria, 698 articles were removed. 41 full-text articles were assessed for eligibility, 39 of these papers were excluded, leaving two full texts for synthesis (see Figure 1). The commonest reason for exclusion was that the service was metropolitan or urban-based.

**Figure 1.**
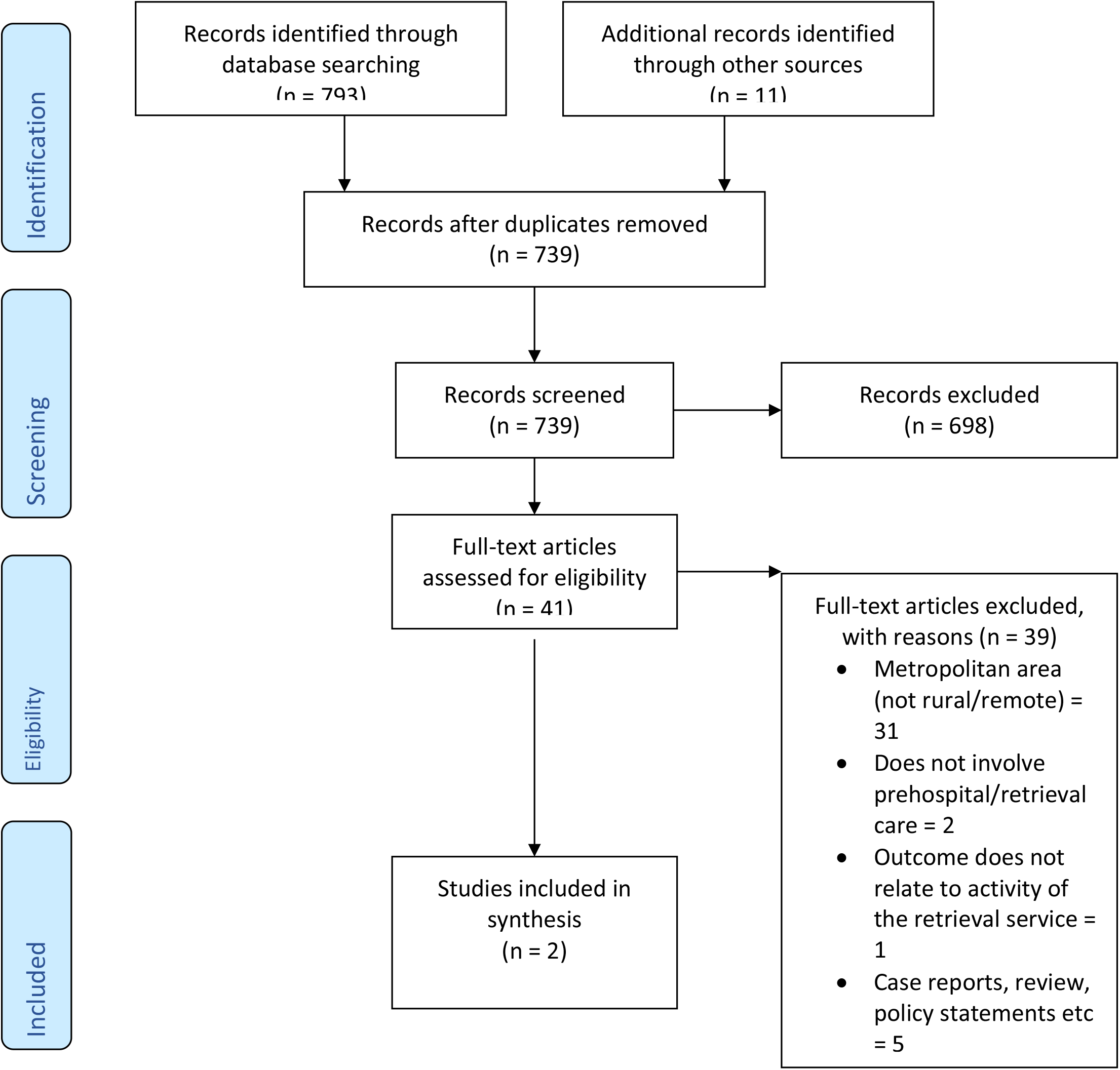
PRISMA Flow diagram of identified, included and excluded studies.

The included papers are shown in Table 2 and were both retrospective observational studies from Australia. Both papers divided the included state into pre-defined regions, based on both rurality and climatic zone, and assessed the impact of heatwaves on these areas separately.

**Table 2.**
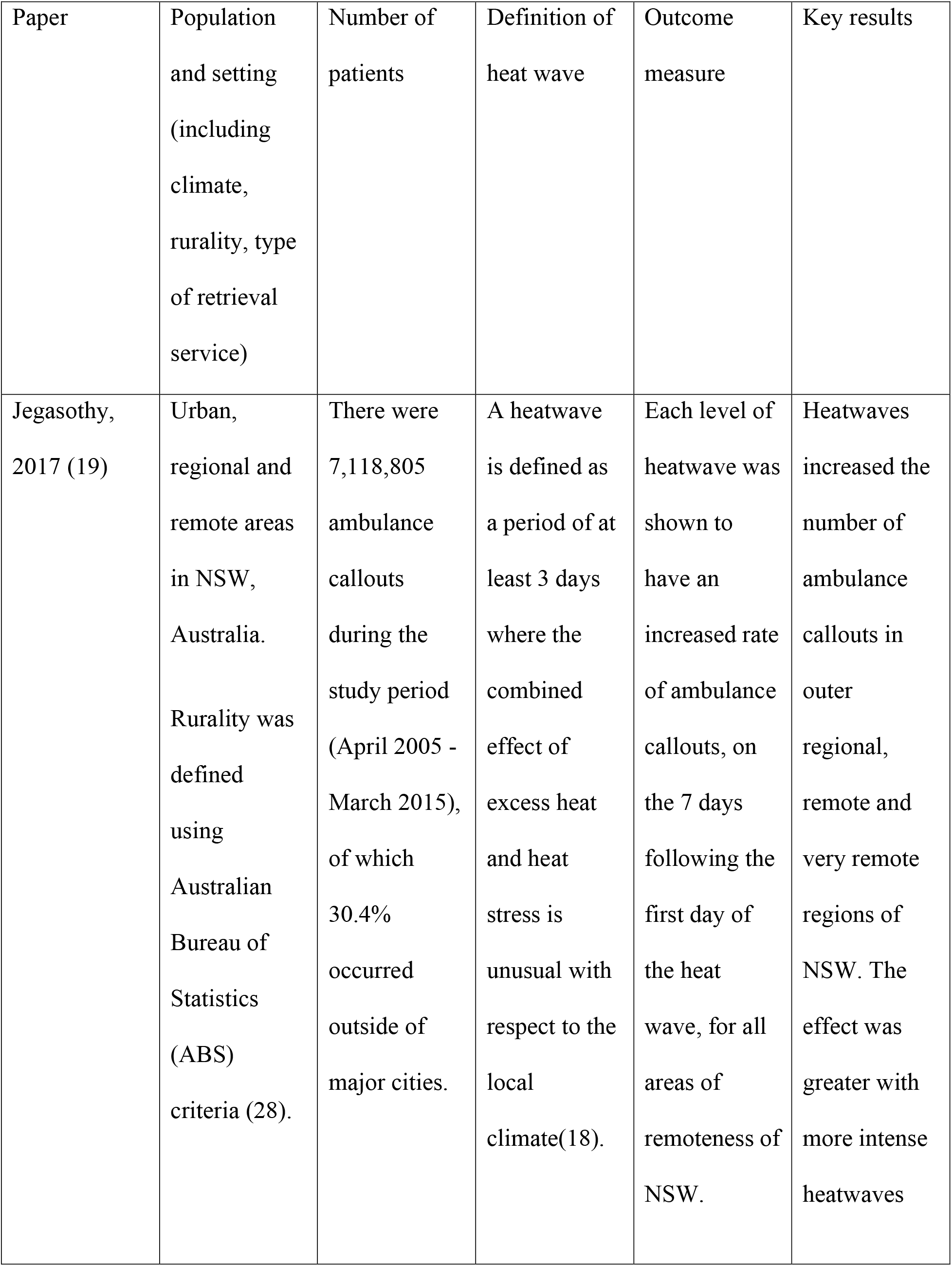
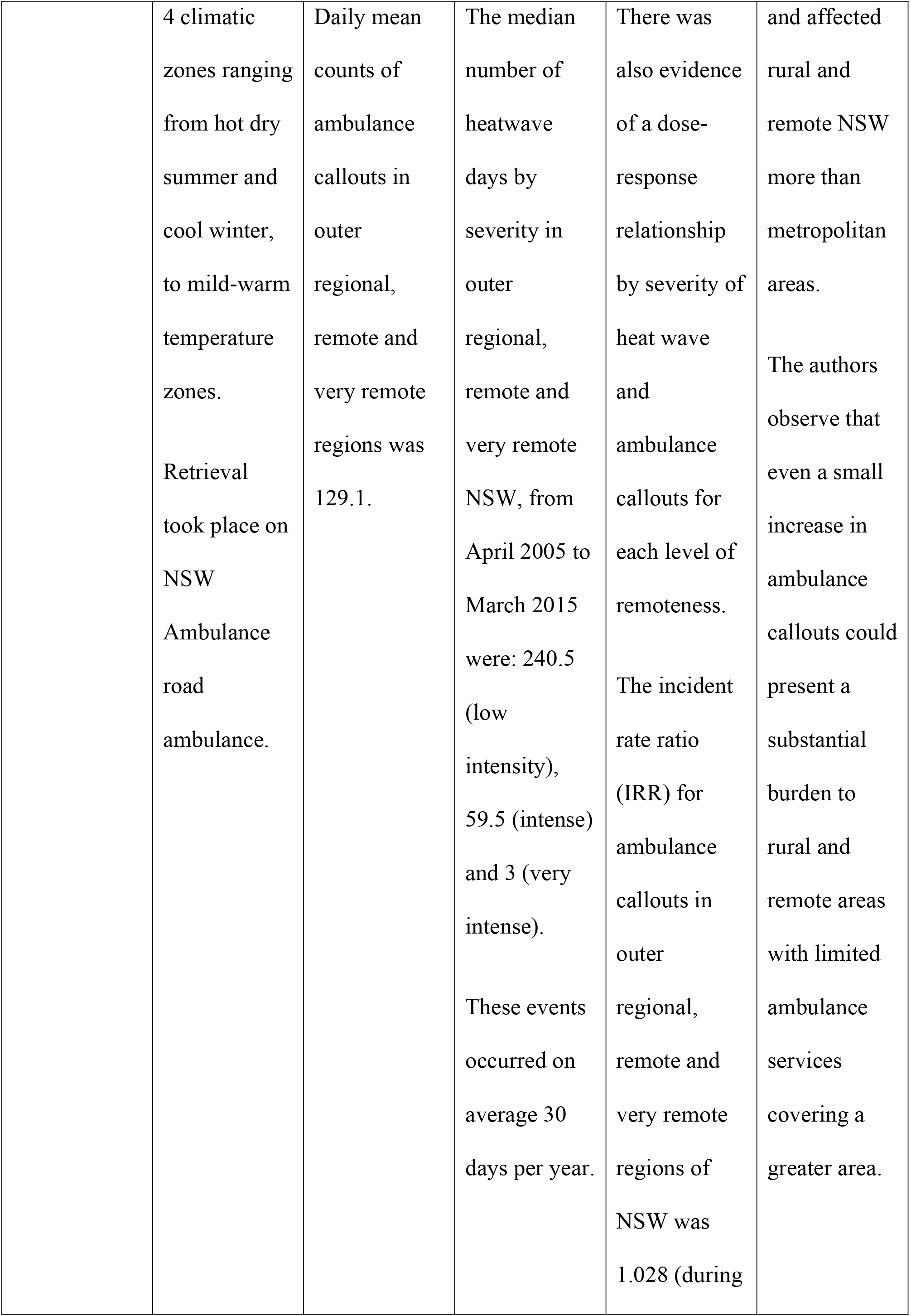
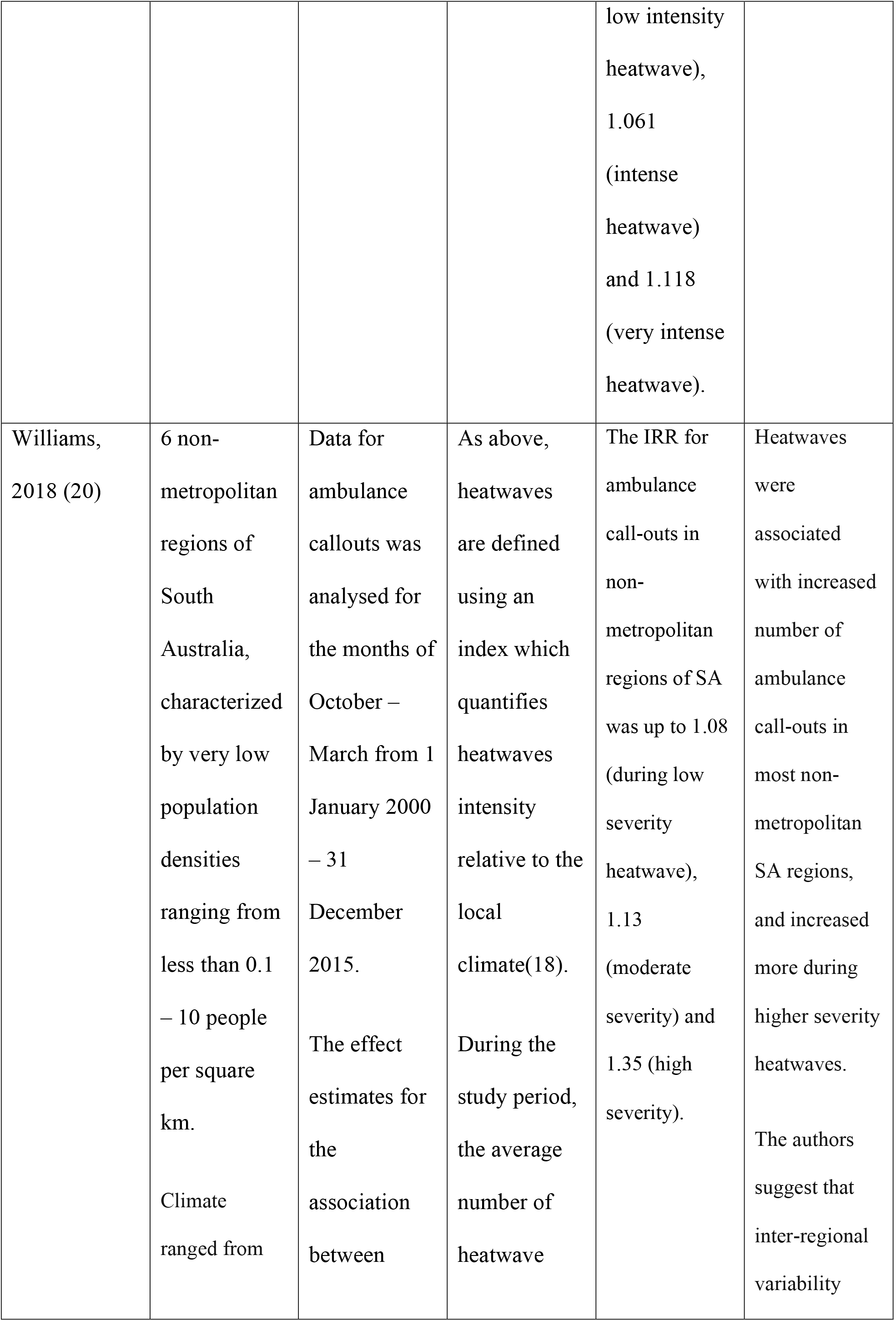
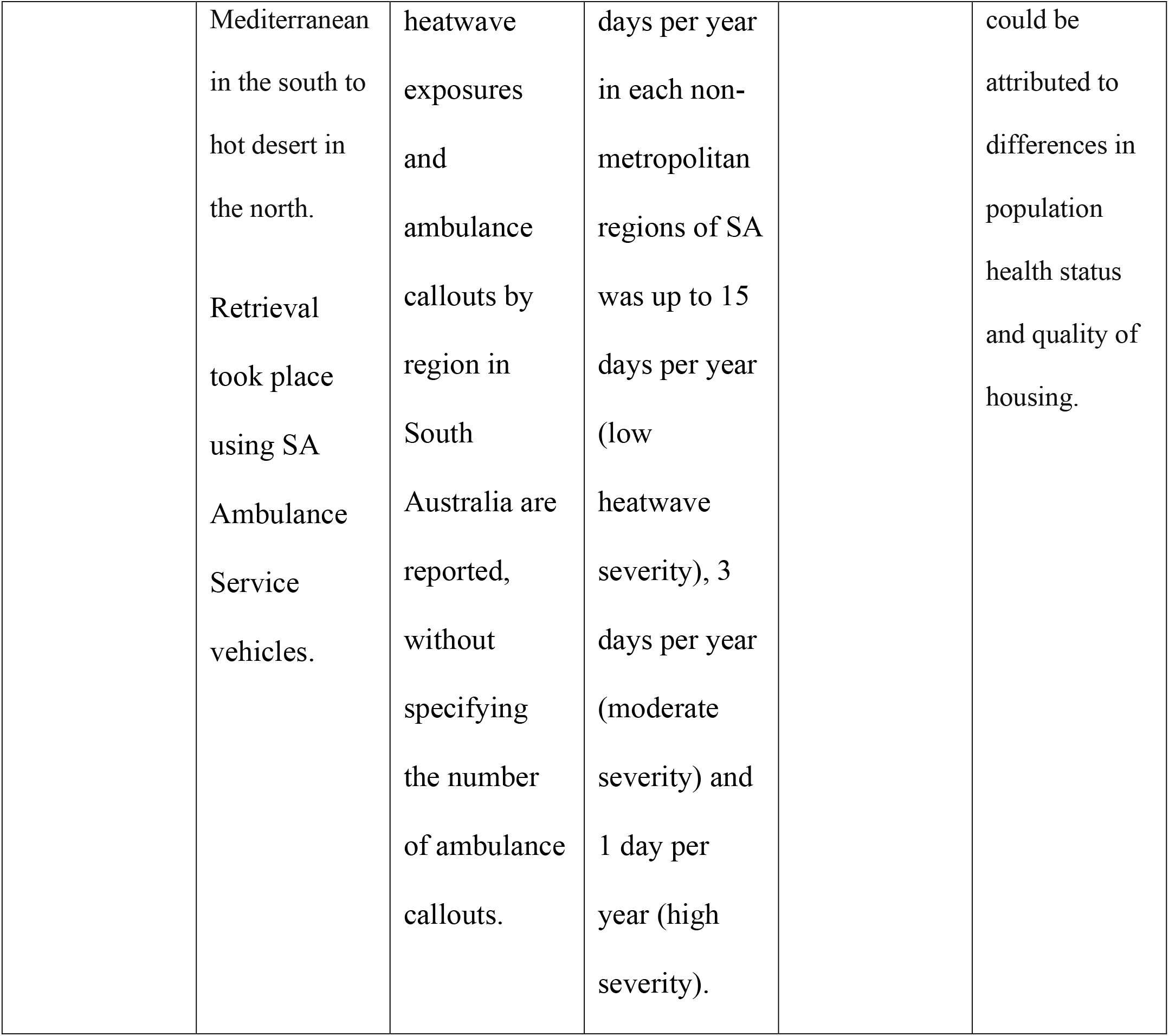
Summary of studies on the impact of heatwaves on prehospital retrieval service activity.

This included urban, regional and remote areas, with climates ranging from temperate coastal regions, to hot, dry, arid zones. Neither study included tropical climates.

The retrieval service platform in both these settings was road ambulance only. There were no reported studies on aeromedical retrieval.

Heatwave was defined in both studies using the Excess Heat Factor (EHF), which is an index based on mean daily temperature data, compared to a local climate norm, as defined the BoM (18). These were subdivided into three different levels of heatwave severity based on the ratio between the EHF and the historical 85^th^ percentile norms from the local area. These were low (0-1), moderate (1-2) and high (>2) severity. Non-heatwave days were defined as a ratio <0 in both studies.

Both studies showed a statistically significant increase in ambulance call-outs following heatwaves across the state. There was also evidence of a dose-response relationship, with increased effect size seen in high severity events compared to low severity events.

Jegasothy et al.(19) found that heatwaves had an increased effect on the number of ambulance call-outs in outer regional, remote and very remote areas, compared to inner regional and major cities. This difference was most pronounced in the low severity events.

Williams et al.(20) also found regional variation, although the picture was more mixed. The greatest effect of heatwaves on ambulance call-outs were seen in the hotter, dryer, more remote inland areas, whereas the effect size in non-urban coastal regions with temperate climates was smaller and was only statistically significant for the low severity heatwave events, which were the most common events. The urban areas fell into the middle of the spread. The authors postulate that the regional variations may be due to sociodemographic factors.

## Discussion

In this scoping review, we found that there was almost no primary literature on the impact of demand for prehospital and retrieval services specifically in non-urban areas, and no research specifically examining this issue in tropical locations, demonstrating a gap in research that should be addressed. Furthermore, we did not identify any data specifically related to aeromedical transport, with only road ambulance transport platforms represented in our findings. There was no universal definition of rural or remote regions in these studies.

Heatwaves were defined in both studies using the EHF metric, which takes into account the usual temperatures for the region in order to allow for physiological and behavioural adaptation to the local climate. However, it does not account for factors like humidity or the existence of a hot season lasting several months that occurs in tropical regions, which are not identified as heat events using the EHF but nonetheless impact on human health and demand for healthcare services.

There are important implications from the results of the study. Firstly, further research is required on measures of extreme heat in different climatic zones, including the impact of humidity and heat ‘seasons’ that are prominent in tropical locations. Effects of extreme heat should also be explored in relation to area-level socioeconomic status and other indicators of vulnerability, in order to improve our understanding of populations that may be more susceptible to extreme heat events. Economic analysis should also be undertaken to quantify the monetary costs of extreme heat on health services, particularly in locations where retrievals require aeromedical transport and can subsequently be very costly.

As the frequency and intensity of heat events increases with climate change, healthcare services should investigate and implement strategies to adapt to and mitigate the effect of heatwaves.

Adaptive strategies may include urban planning and increasing the availability of infrastructure and resources in rural and remote areas in anticipation of heat-related increase in activity. Economic analysis may demonstrate that the benefits of improving housing quality in remote Australia outweighs the costs of initial investment. Retrieval services may need to consider occupational health interventions to keep clinicians and crew safe when working during extreme heat (21).

Mitigation strategies could include advocacy within healthcare to reduce the environmental impact (22, 23), given the healthcare sector in Australia is responsible for 7% of Australia’s total carbon emissions(24). Failure to invest in adaptive and mitigation interventions is predicted to increase the demand for aeromedical retrievals, which in turn is likely to increase carbon emissions.

### Limitations

This review only included English language publications. Both studies were from Australia so the relevance to other countries is unknown. No risk of bias assessment was performed therefore it is not possible to assess the quality of the evidence-base.

### Conclusions and recommendations

It is well established that heatwaves increase demand for emergency transport in metropolitan areas (4, 25-27), however this review found almost no primary literature on demand for prehospital retrieval services in rural and remote areas, and no data specifically related to aeromedical transport. The research did recognise the disproportionate impact of heat-related increased service demand on rural and regional health services.

There is an urgent need for more research in this area, with the impact of climate change already being felt amongst vulnerable residents of remote communities in Australia. The results of this literature review have informed an observational study of the impact on heatwaves on a remote Australian retrieval service activity to quantify this effect. This data is vitally important in supporting advocacy for mitigation and adaptations strategies.

## Data Availability

The data that support the findings of this study are available from the corresponding author upon reasonable request.

## Acknowledgments

There was no external funding for this work and researchers provided in-kind resources to the project.

